# Cumulative Active and Recovery Rates Based Criterion for Gradual Lockdown Exit: A Global Observation of SARS Cov-2 Management

**DOI:** 10.1101/2020.06.05.20123364

**Authors:** Dhananjay V Raje, Abhay Bajaj, Moumita Chakraborty, Hemant J Purohit

## Abstract

On March 11, 2020 the World Health Organization (WHO) had declared SARS-CoV-2 pandemic; and at present there are over 6 million cases across the globe. Based on the interim guidelines of WHO, most of the countries opted for social distancing with lockdown as the only way to control the pandemic. This led to ‘Manufacturing’ shut down, which acted as a spanner in the wheel for international supply chain leading to pressure on governments to review the protocols of the lockdown. We studied epidemiological parameters for 18 countries and obtained crossover time point referring to cumulative case active and case recovery rates and the time point for the peak positive confirmation rate in a time window of 92 days; and linked with the respective governmental decisions. For countries awaiting crossover, time series non-linear models could be used for predicting the crossover point. A sample study was carried out for India. The median time for reaching crossover for 12 countries was 37 days, while peak positive confirmation rate was 30 days after their first intervention. These countries enforced strict lockdown regulations and have shown constant improvement in their recovery rate even after crossover time point. A phase wise relaxation of lockdown is evident after crossover point in most of these countries. The crossover time point with the subsequent increasing recovery rate can be a strategy for lockdown relaxation as evident from the experiences of few countries. Also, we propose a criterion based on 28 cumulative recovery and fatality rate for micro-management of lockdown.

## Introduction

World economy is a network of the global value chain, where every single country is a partner. By March 2020, the whole civilization realized the pandemic impact of SARS CoV2; and world over, the restrictions in movement and lockdown have severely impacted the industrial supply chain.^1,2^ The cumulative effect of this interrupted productivity led to loss of millions of jobs and this has now created uneasiness at the different levels of society; and hence, governments are looking for solutions to come out of this lockdown situation.^3–5^ Different modelling approaches have been reported to predict the course of the viral spread, of which susceptible-exposed-infected-recovered (SEIR) model, is the most explored approach applied by various groups with data collected from different countries.^6,7^ However, due to the unpredictable propensity of this virus and the sporadic identifications of surge clusters in the society, there were interim directives made by different governments. Inclusion of such inputs limits the prediction accuracy of model for the course of a pandemic.^8–10^Another issue is the diversity as reflected by genome sequence data that suggests, virus is either mutating or adapting according to host in different part of the world.^11^.

In this paper, we have opted an observational approach using the data for monitoring test with active, recovered and fatal cases and whenever the model showed some perturbations, we have tried to link with the government interventions, which could have influenced the spread of the virus through usual or community transmission mode.

## Method

In this study, an observational approach was adopted in which epidemiological parameters were observed with time and retrospectively analysed with reference to interventions or public announcements by the governments.

Eighteen countries mostly from Europe and Asia were considered with at least 1000 diagnosed cases as on March 31^st^ 2020. All these countries were tracked from March 1^st^ for the number of new cases, active cases, recoveries and number of fatalities. The data source was *John Hopkins CSSEDISandData*, available as an open source. This data was used to derive the case active rate (CAR), recovery rate (CRR) and the fatality rate (CFR) at each discrete time point till May 31, 2020. These rates were obtained as the cumulative number of active/recovered/fatalities on a specific day to the total number of cases diagnosed till that day, considering a lag of zero days. Thus, on any day, the total infected individuals were categorized into these three states, summing to unity. Moreover, the ratio of number of newly detected cases to the total number of tests performed was also obtained giving the positive confirmation rate (PCR) of the disease with time. The data source for the number of tests was ourworldindata.org.^12^ All these rates were plotted in the same frame with a time window spanning 92 days i.e. from March 01, 2020 to May 31, 2020. The interventions in the form of national recommendation, lockdowns in each of these countries during this period were linked with the progression of CAR, CRR, CFR and PCR. The foremost interest was to observe the crossover time point of active and recovery rates for each country. This is the time point at which the CRR is either greater than or equal to CAR. Also, the time point for peak PCR i.e. the time at which the PCR attains maximum, was observed for each country.

For countries in which the crossover point has not reached, time-series non-linear models could be used for prediction. A sample study was performed for India in which piece-wise linear regression and quadratic regression models were fitted to predict the crossover point at the end of 60^th^ and 75^th^ day in the lockdown period. The residuals were obtained to decide how well the models capture the trend.

Further, a *hierarchical* strategy of disease management has been proposed for multistate countries, with each state comprises of cities/districts. Two outcome criteria i.e. CRR and CFR can be referred to indicate the COVID19 status of each city/district within the state. The CRR at the crossover time point and the corresponding CFR at the same time point can be used to determine the hot-spot cities/districts within the state. A sample study was performed for India, in which the most affected states were mapped based on the above rates. Further, a sample exercise was performed for Maharashtra state, wherein the cities/districts were mapped using the above rates. Thus, the city/district level information is available for state government, while state level information is available to central government for necessary interventions.

## Results

Among the selected countries, seven were from Asia, while eleven were from European continent including Russia, which is transcontinental. The CAR, CRR and CFR for each day along with the PCR were obtained for each country. Simultaneously, the major events and interventions relevant to the disease dynamics were studied for each country. The trends for twelve countries attaining the crossover are shown in Figure 1 along with the government interventions.

**Figure 1:**
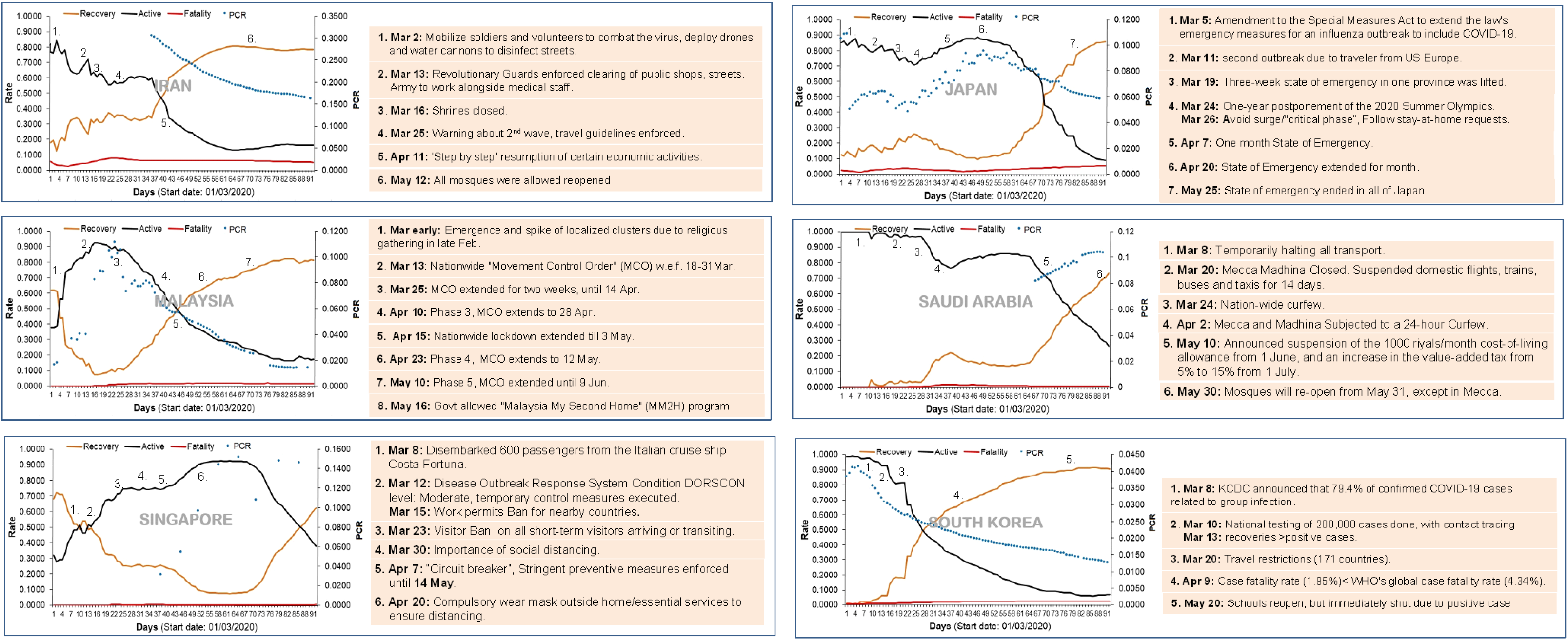
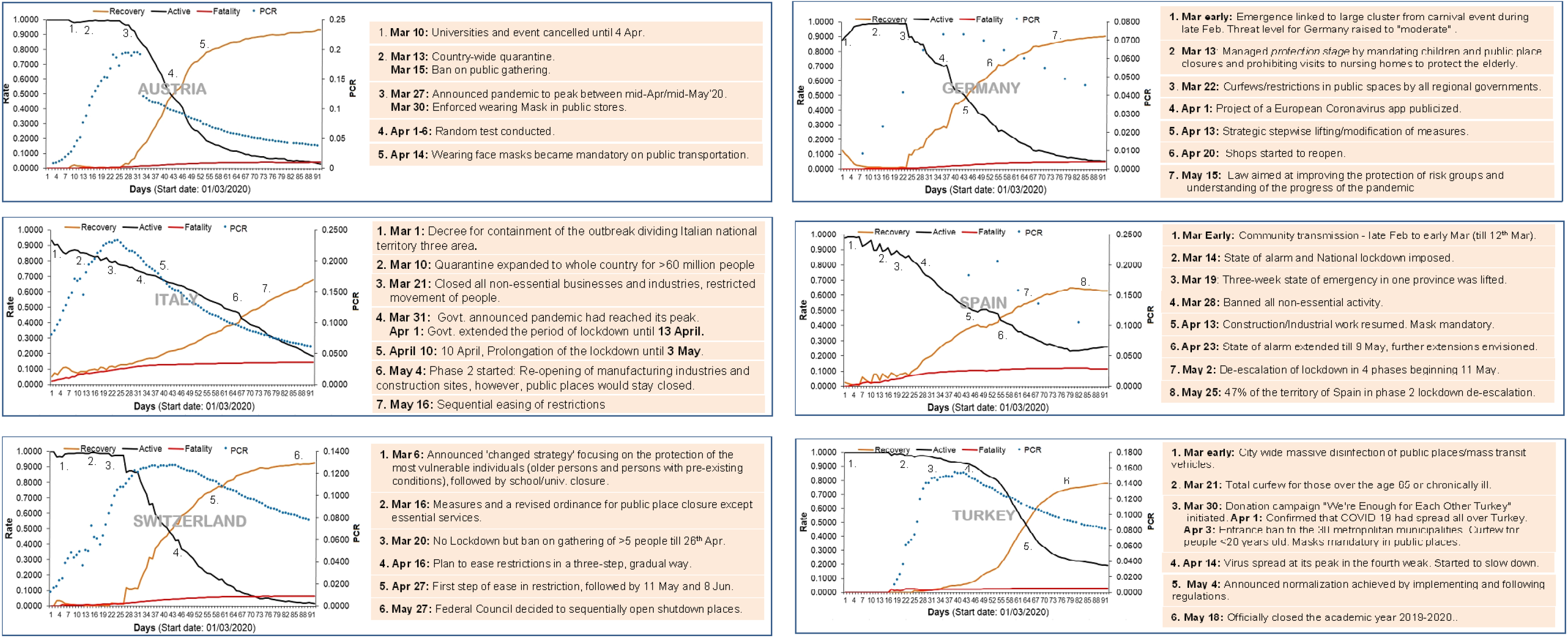
The epidemiological measures for Asian (A) and European (B) countries achieving crossover time point within the observation window of 92 days starting from March 01 – May 31, 2020. Alongside is the description of various events and interventions by the respective governments before and after crossover time. It seems that in eight of these twelve countries, the decision for relaxation of lockdown has been taken after reaching the crossover time and deceleration of PCR. A prior peak for PCR followed by a crossover could be a controlled scenario for any country to manage COVID19 pandemic. Subsequent to crossover, the CAR and CRR were found progressing in the same acquired directions in logarithmic manner. Based on these trends, the strategies by these governments appears to be rational, and can be adopted by countries in which the crossover is still awaited.

In *Austria*, the PCR increased exponentially till the end of March. By this time, the CAR and CRR were almost constant and the CFR was negligible (< 1%). The effect of intervention surfaced by the end of March, where the PCR reached a saturation level and subsequently started declining. The CAR and CRR deflected by March end and showed a crossover on April 12^th^ with CRR (0.5010) exceeding CAR (0.4739). The CFR was 2.4% at the crossover. Post-crossover, the situation in the country started improving with 93.2% recovery and 3.99% active cases as on May 31^th^. The time of peak PCR occurred nearly 12 days before the crossover point in Austria.

In *Germany*, the PCR increased across country till March end, while the CAR, CRR and CFR were constant. After 22^nd^, the CRR gradually headed upwards, and by April 13^th^, the country attained the crossover with CRR (0.4943) marginally exceeding CAR (0.4811). Moreover, the PCR attained the peak by the same time, which subsequently showed a decline. Post-crossover, the government adopted strategies for stepwise modifying the measures against pandemic. The country has 90.1% recovery and 5.19% active cases as on May 31^st^.

In *Italy*, the PCR was steep from March 1^st^ to March 25^rd^, and later showed a consistent decline. The crossover was achieved on May 6^th^ with CRR (0.4348) marginally above CAR (0.4268). The peak PCR reached by 21^st^ March and occurred nearly 45 days before the crossover. Post-crossover, the CRR is continually increasing with government restrictions with 67.6% recovery and 18% active cases as on May 31^st^.

In *Spain*, the CAR and CRR showed a gradual change with high fluctuations till March 21^st^. Although the PCR was increasing, the CRR showed upward trend with time. The crossover was achieved on April 24^th^ with CAR (0.455) exceeding the CAR (0.4341). The time of peak PCR almost coincided with the crossover. By 31^st^ May, the country had 62.8% recovery and 25.8% active cases.

In *Switzerland*, the PCR increased up to April 5^th^, while CAR, CRR and CFR were almost constant till March 25^th^. A crossover was reached on April 12^th^ with CRR (0.4997) exceeding the CAR (0.4568). The PCR started declining after April 12^th^. The crossover and the time point of peak PCR coincided for Switzerland. Post-cross over, the CRR increased consistently and reached to 92.3%, while active cases were only 1.4% by May 31^st^.

In *Turkey*, the spreading peak of virus was noted on April 12^th^, and subsequently, a gradual slowdown was observed till May 15^th^. The CAR started declining after April 23^rd^ and the crossover reached on May 3^rd^ with CRR (0.5010) exceeding CAR (0.4720), and CFR around 2.7% continuing till the end of observation period. The time of peak PCR occurred 21 days before the crossover point in Turkey. By 31^st^ May, the recovery was 78.1%, while active cases were 19.2%.

Among Asian countries, in *Iran*, the CAR and CRR showed fluctuations with gradual decline from March 1^st^ to 25^th^. On April 9^th^, a crossover was observed with CRR (0.4879) exceeding the CAR (0.45). The CRR increased logarithmically till May 1^st^ and remained constant even after relaxation. The positive confirmation data available from March 5^th^ showed an exponential decline till May 15^th^. As on 31^st^ May, the country has 78.5% recovery and 16.4% active cases.

In *Japan*, till March 25^th^, there was almost a constant PCR of around 5%, with the decrease in CAR and corresponding increase in the CRR with CFR near to 3.4%. The PCR and CAR were at peak on April 19^th^, which later showed gradual declines. The crossover reached on May 10^th^ with CRR (0.5151) exceeding CAR (0.4453). After May 10^th^, the CRR continued to increase, while the CFR gradually increased to 4.4% by the end of the observation period. At the end of observation period, the country had 85.6% recovery, while 9.02% active case.

In *Malaysia*, the spread of virus increased sharply from March 1^st^ till 23^rd^. The CAR also showed increase till this time point with a corresponding decrease in the CRR. The crossover reached on April 14^th^ with the CRR (0.4969) exceeding CAR (0.4867). Post-crossover, the CRR continued increasing till the end of observation period. The time of peak PCR reached 21 days before the crossover. The recovery was 81.2% and active cases were 17.3% by May 31^st^.

In *South Korea*, the spread of virus reached a peak on March 3^rd^ and thereafter showed a gradual decline by the end of May. The CAR and CRR were almost constant till March 10^th^; however, subsequently, there was a sharp change in these rates resulting into a crossover by March 28^th^ with the CRR (0.5076) exceeding CAR (0.4772). Post-crossover, the trends continued along with reducing spread as on May 31^st^. The time of peak PCR reached 25 days before the crossover point.

In *Singapore*, the CAR increased till April 22^nd^, and then was constant till May 9^th^. Later it started declining and by May 27^th^ provided the crossover. Subsequent to this, the CAR continued declining with increasing CRR. The peak PCR was observed by May 4^th^ and was constant till the end of May. The recovery was 62.2%, which active cases were 37.8% by the end of observation period.

In Saudi Arabia, the CAR and CRR were almost constant till March end. There was a deflection in the trend, which continued till April 10^th^. Again, there was almost a constant phase for both the rates till May 3^rd^. Later, there was consistent decline in CAR and increase in CRR with a crossover on May 20^th^. Post-cross over the trends continued with a recovery of 73.2% and active rate of 26.2% as on May 31^st^.

For other countries like *France, India, Russia, Sweden* and *United Kingdom*, the crossover could not be observed till the end of May, despite different measures by the local governments (Figure 2). The crossover point could be predicted for such countries using appropriate linear/non-linear models. The analysis for India have been presented.

**Figure 2:**
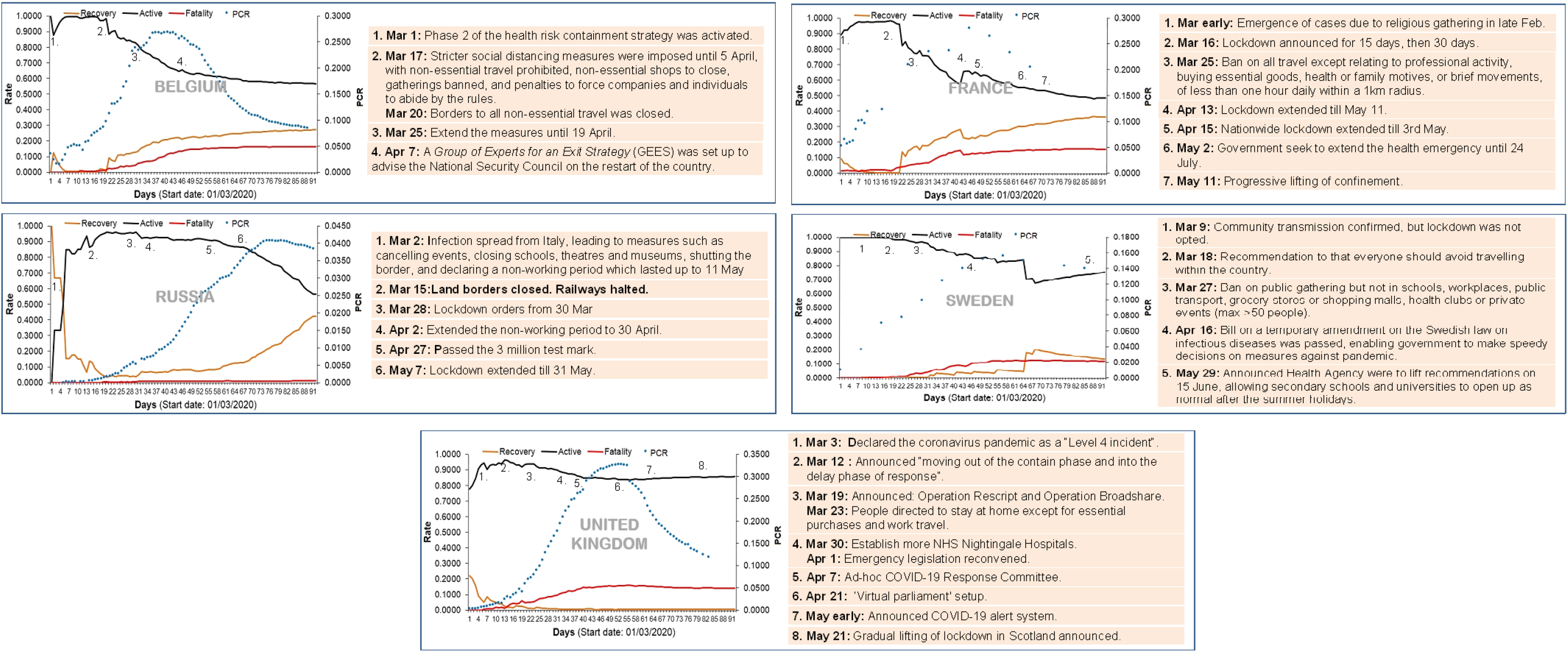
The countries from Asia and Europe with crossover awaited. The adjacent panel provides the description of events and government interventions till May 31^st^. These nations have either attained the peak PCR or still awaiting.

## Cross-over forecasting – A study for India

In India, since March, the incidence of COVID19 is on its rise. Figure 3 provides the CAR, CRR and CFR, which shows that the crossover point is very close but not reached in the observation window. The PCR increased till April 6^th^ and later continued in the range of 4 –4.71% till May 15^th^. However, subsequently, the rate is increasing exponentially and has touched 5.1% by May 31^st^. Since April 10^th^, the CRR started increasing; nevertheless, the crossover is still awaited. The fatality rate is around 3% since April 1^st^.

**Figure 3:**
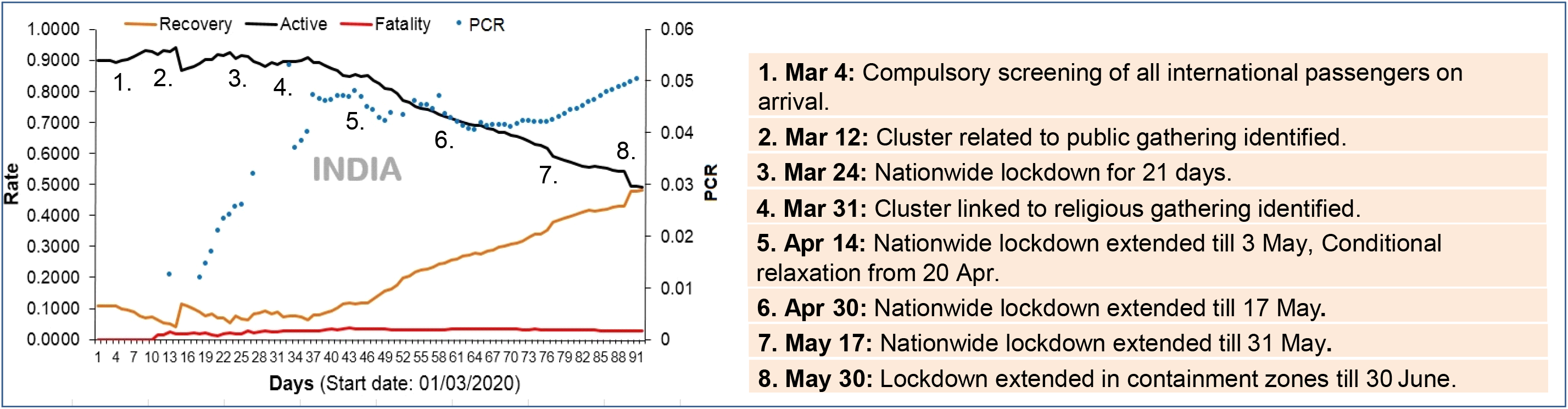
The trends of epidemiological parameters for India in the observation window of 92 days. The adjacent panel provides the description of events and government intervention from March 20 onward. The crossover for the country is yet awaited. India is the only country in the study list showing a prolonged stationary PCR till May 15^th^ and thereafter an exponential increase. The CAR and CRR are indicating a crossover by May end, provided the CFR remains stable around 3%.

A predictive modelling to estimate the crossover time point was carried out using Piecewise linear and quadratic models (Figure 4A-B), which shows graphical visualization of model fits along with predictions for piecewise model with 80% and 95% confidence bands. The predictions were done after 60 days and 75 days with March 1^st^ as zero^th^ day. For piecewise model, a single knot at *c* = 40 days was considered for fitting purpose. Based on 60 days (Figure 4A), the quadratic model predicted the crossover on 72^nd^ day, while the piecewise model predicted on 84^th^ day. The analysis based on 75 days provided crossover on 90^th^ day by quadratic model, while 91^st^ day using piecewise model. Thus, it is evident that the piecewise regression model was relatively more consistent in predicting the crossover irrespective of the training set. From April 10^th^, the two rates are following almost a linear trend. The predictions by quadratic model deviated drastically with 60 days data sets, while overlapped considerably with piecewise model for 75 days data set. The heteroscedasticity in the rates was less after 40 days as evident from the figure, resulting in minimal residual error for piecewise regression model. So, with continuing restrictions, the crossover was expected by 91 days i.e. end of May for India; however, could not be achieved when observed with actual data. Tentatively, now expected in first week of June.

**Figure 4A-B:**
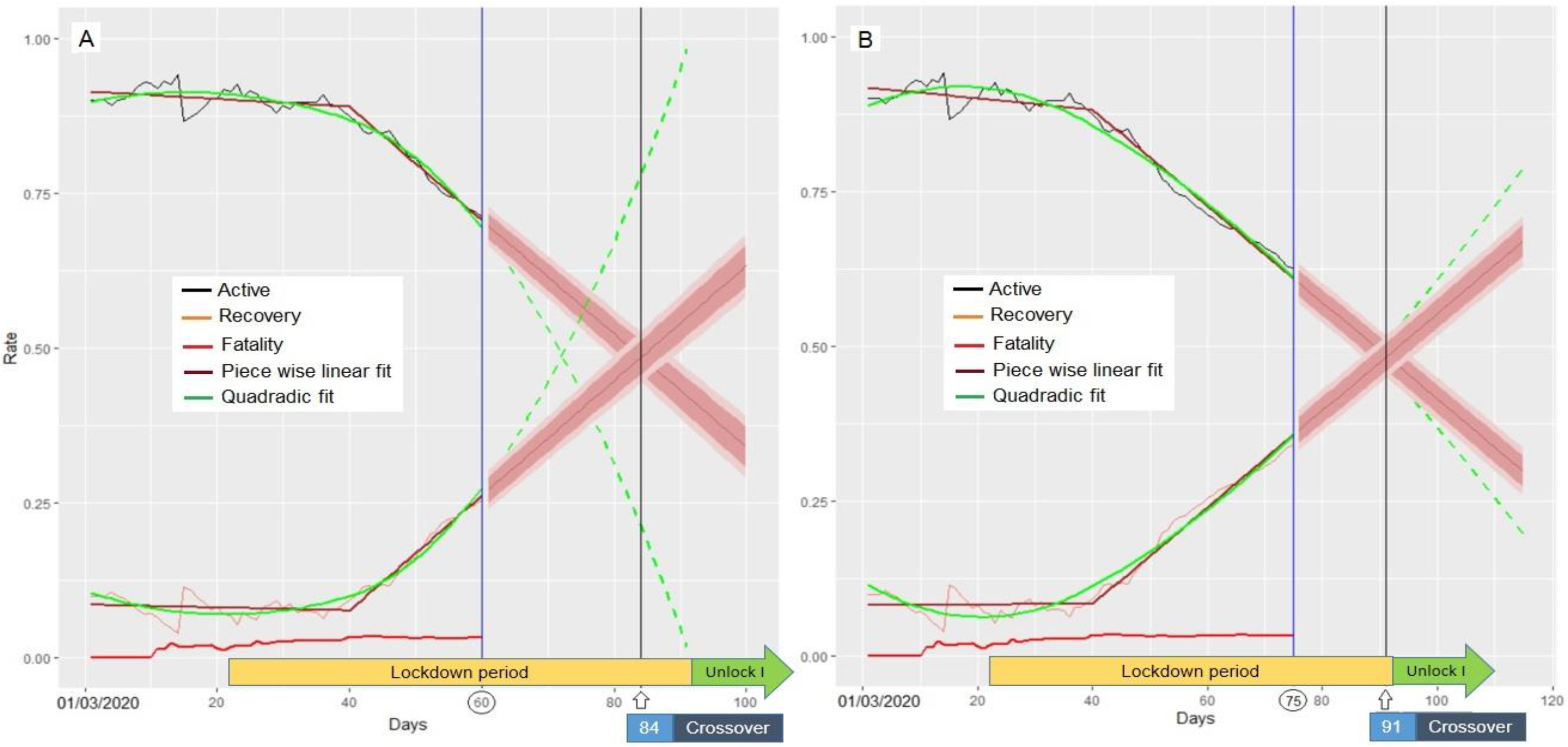
The active, recovery and case fatality rates at different days of COVID19 progression starting from March 01, 2020 in India along with predictions. A) Model predictions for active and recovery rates based on 60 days B) Predictions based on 75 days. The 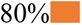 and 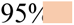 confidence intervals are shown for Piece wise linear fit. The cross over point is the time (in days) of intersection of active and recovery rates.

Since, it was observed that once a country crosses 50% cumulative recovery cases, the trend is always followed. Hence, we first predicted the possible time point for 50% cumulative recovery of cases for India; as described earlier. The next step was how to derive a criterion, which can propose the strategy to gradually exit the lockdown with national data supporting 0.5 CRR. We have applied following disease management strategy, we propose two parameters viz., CFR and CRR that can be jointly assessed for each state and eventually each city/district. The thresholds for these parameters can be referred based on overall national data for CRR and CFR; and accordingly, the classification can be obtained at each hierarchical level. Figure 5A provides the country level representation in which different states (majorly affected) have been positioned as per the CFR and CRR status on May 31^st^. The states in each quadrant can be further assessed at next hierarchical level i.e. cities/districts. Figure 5B gives the state level representation for Maharashtra. The hot-spots within the states and their movements with time can be observed and accordingly the action plan at city level can be determined. Figure 5C provides the plans for the four zones, which are subject to alterations.

**Figure 5A-C:**
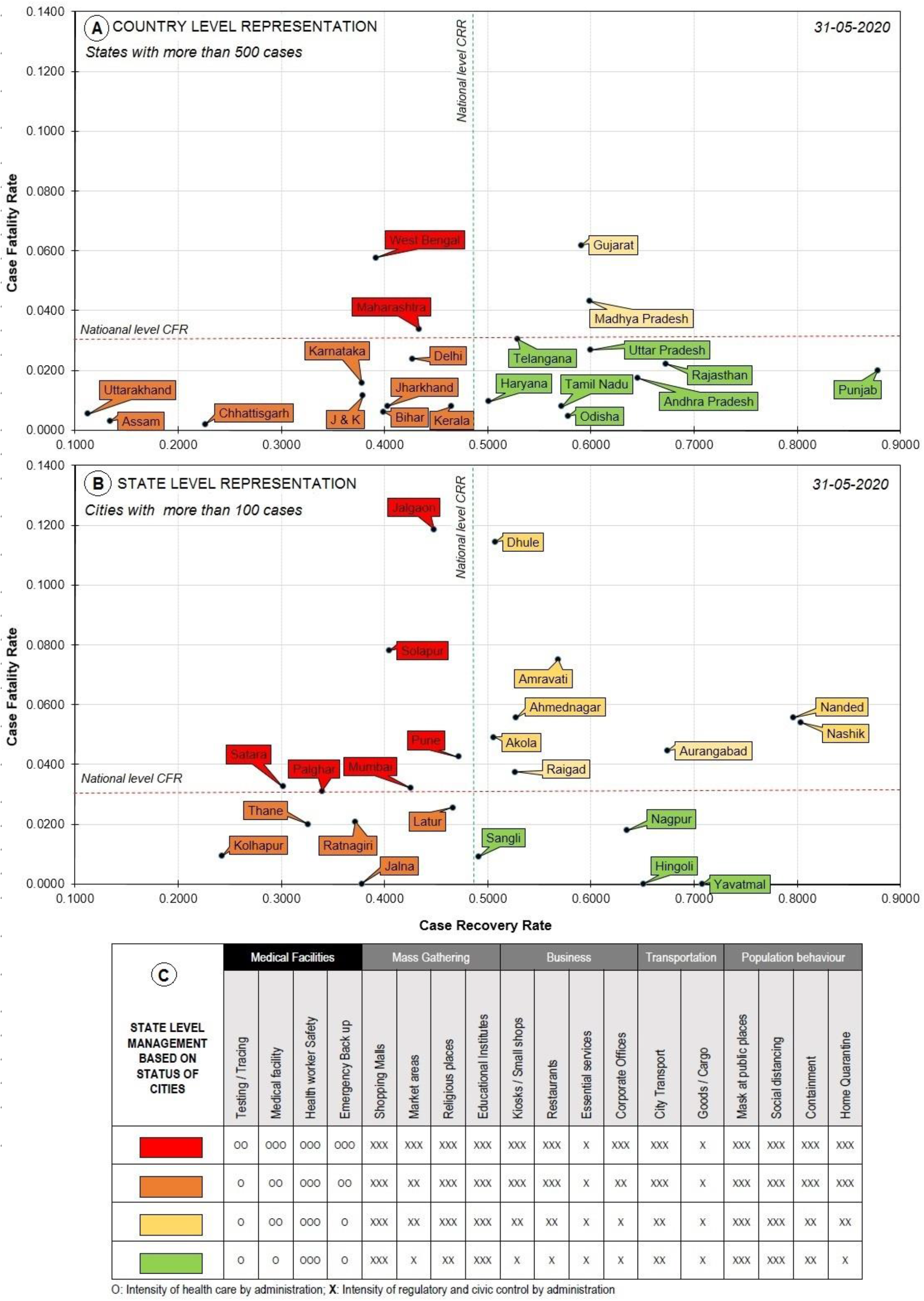
The 2D representation based on CFR and CRR in different quadrants. A) Country level representation for India showing states with more than 500 cases B) State level representation for Maharashtra state showing cities with more than 100 cases as on May 31^st^ C) A tentative state level management guideline to improve upon the two parameters. The dashed lines indicate the national level CFR and CRR as reference to identify the hot-spots.

Since, the disease progression is dynamic, it is essential to track the movement of each city and thereby the state on a daily basis. A dashboard has been prepared, which at present provides the updates for the most vulnerable state i.e. Maharashtra, which has ∼34% of the active cases of the country. We are expanding this platform for the critical states of the country, so that the day wise state level information is available for all critical states. The dashboard can be viewed at: https://covid19stat.shinyapps.io/covid19status/.

## Discussion

The issue with coming out of lockdown or relaxing scenario in between different periods of lockdown is an unexpected surge of transmission waves.^13–15^ It challenges the medical infrastructure with overburdened staff and stressed supplies; and this seems to be true for almost every country.^16–18^ The volatility analysis of wold economy suggests there will be a deep depression around the world in the year 2020;^3,19,20^ and WHO recently stated the virus will be around for a while. In such scenario, every country has to delineate the strategic time lines to exit lockdown, since it is crucial to balance the health and economic burdens.^21–23^

In this study, we adopted a strategy of learning from the experiences of select countries based on epidemiological parameters. The primary focus was on crossover time and the time of peak PCR. The government interventions were linked with these two parameters. The analysis of twelve countries revealed that the two time parameters have association with the governmental decisions, as evident from Figure 1; and that the spread of SARS-CoV-2 can be effectively modulated. In countries where initial lockdown measures were strictly implemented, like home quarantine, ban on public gatherings, conditional movement only for essential services, and closing of education system, the PCR could be controlled and the crossover could be achieved. The median time for reaching crossover for 12 countries was 37 days, while median peak PCR time was 30 days after their first intervention. In six out of the twelve countries, the crossover time occurred after time of peak PCR with a median of 21 days; while in three countries i.e. *Germany, Spain* and *Switzerland*, the times for two parameters coincided. Further, post-crossover, it was observed that in these countries, with step-wise relaxation of lockdown, the CFR and CRR continued in the acquired directions indicating better pandemic control. The countries like *Belgium, France, India, Russia* and *United Kingdom* (UK) are awaiting the crossover point or the peak PCR. In *Belgium, France* and UK, the crossover is much awaited considering the CAR, CRR and CFR trends. The patients are under prolonged treatment and thus confining the recovery rate. Perhaps age could be one of the factors, as the population median age of these countries is above 40 years. In *India* and *Russia*, the crossovers are nearby. It would be interesting to observe the progression of two rates in these countries; and especially for India, which is stepping in the unlock phase I. During the lockdown period, the country has seen improvement in the CRR with controlled CFR; however, could not reach the crossover. Based on the experiences of other countries regarding lockdown and relaxations, it is anticipated that the two rates should continue with the acquired trends, and that the current CFR remains steady and PCR shows deceleration. To achieve this, the unlocking phase needs a careful monitoring of cities and in turn the states. The hot-spots will depend on the current national CFR and CRR rates, which are dynamic with time. The proposed real time dashboard could be of immense utility at these moments.

Another interesting observation comes from the genome sequence data of virus, with over 32000 genome sequence available globally (https://www.gisaid.org/epiflu-applications/nexthcov-19-app/). The data suggests that there are seven major clades which can group most of the sequences with every clade having significant variation at the individual sequence level. There is significant variation in sequences after mid-February. The viral sequences can be grouped in two different clades which are associated with European sequence data. Whereas, Asian sequences are close to two major European clades, with an additional clade having different sequences representing different countries.

There is a need to understand the whether the strains/sequence data observed in the twelve countries have any association with the peak PCR and the crossover times observed in these countries. This may reveal the relevance of different strains and its interaction with host machinery, which is supporting the viral multiplication and its dispersion factor.^24^ The viral genome sequence data analysis from India shows two major clades.^25,26^ However, the sequence data is not having enough representative sequences from the country, and hence cannot be correlated with number of active cases observed in different states for the moment.

For some countries, the crossover point is before 50 days, while for others it is beyond. It will be interesting to map the sequence data from different countries before and after the cross over time, where 50% recovery rate has been achieved. This can further be analysed with countries showing delayed or no crossover point. This pattern of dispersion of infection needs further analysis with its correlation with demographic or even with data from the virus-host interaction studies.^27^ Irrespective of genetic property of virus and its multiplication in host, it’s evident from the analysis of different countries that the regulation imposed as lockdown and restriction in activities has real role in controlling of this pandemic. The other important observation emerged that once a country crosses 50% cumulative case recovery, then it always follows the improvement trend, even though in cases of few countries, there were surges of positive cases around the cross over point was observed.

## Conclusion

The exit from lockdown is the only imperative option for any economy to come back to its normalcy, and that means the restoration of supply chain so that every component of society gradually recovers from scarcity of resources. The parameters used in this study, like crude active rate, recovery rate and positive confirmation rate are the ‘real indicators’, which are inclusive of many factors like population location, susceptibility, incidences, and surveillance for respiratory infections, which may independently and severely impact the forecasting. By referring to the criteria of crossover and peak time point of positive confirmation rate, policy makers can assess the impact of the decisions being taken up. This proposal advocates relaxing norms for state or district where these criteria are fulfilled but puts the condition of isolating the areas, which are still struggling with unfilled threshold parameters.

## Data Availability

All data referred to in the manuscript is available in public domain.

https://www.gisaid.org/epiflu-applications/next-hcov-19-app/

https://gisanddata.maps.arcgis.com/apps/opsdashboard/index.html#/bda7594740fd40299423467b48e9ecf6

https://covid19stat.shinyapps.io/covid19status/

## Acknowledgement

Authors are thankful to the Director, CSIR**-**National Environmental Engineering Research Institute (CSIR-NEERI) for his kind support. The manuscript has been duly checked and assigned KRC No. CSIR-NEERI/KRC/2020/MAY/EBGD/2.

## Author Contributions

HJP, DVR, AB planned the study. DVR, AB, MC, HJP performed analysis. MC developed the site using R-Shiny App platform, DVR, HJP, AB wrote the manuscript. All authors proof read the manuscript.

